# Cardiovascular Outcomes in Patients with Type 2 Diabetes and Heart Failure with Reduced Ejection using Glucagon-like Peptide-1 Receptor Agonists

**DOI:** 10.64898/2026.09.21.26363576

**Authors:** Mohammed Al-Nusair, Aneesha Maini, Shriya Khurana, Jowan Al-Nusair

## Abstract

**Background:** Glucagon-like peptide-1 receptor agonists (GLP1-RA) improve cardiovascular outcomes in patients with type 2 diabetes (T2D) and obesity, and more recently they have been shown to be beneficial in patients with obesity and heart failure with preserved ejection fraction. Their role in patients with heart failure and reduced ejection fraction (HFrEF) and T2D remains underexplored.

**Methods:** A retrospective cohort study was conducted using data pooled from the TriNetX platform. Patients with T2D and HFrEF who were initiated on GLP1-RA therapy during the period May-13-2026 to Jun-6-2026 were identified. One:one propensity-score matching with controls was performed. Time-to-event analysis was conducted to compare the one-year incidences of all-cause mortality and a primary composite outcome of acute myocardial infarction, ischemic stroke and acute heart failure, between the two matched-cohorts. Secondary outcomes included individual components of the primary composite outcome and any inhospital encounter.

**Results:** Two well-matched cohorts with 38,879 patients in each were formed. Patients treated with GLP1-RAs had a lower incidence of all-cause mortality (4.2% vs. 12%; absolute risk difference [ARD] -7.8, 95% CI -8.17 to -7.41; HR 0.4, 95% CI 0.34─0.38) and the primary composite outcome (32.5% vs. 48.3%; ARD -15.8, 95% CI -16.47 to -15.11; HR 0.6, 95% CI 0.57─0.60) compared to the control group. The GLP1-RA group were also at lower risk of all secondary outcomes compared to the control group.

**Conclusion:** In a real-world study, GLP1-RA therapy was associated with lower risk of all-cause mortality and cardiovascular outcomes in patients with T2D and HFrEF.

## INTRODUCTION

Type 2 diabetes mellitus (T2D) and heart failure (HF) frequently coexist and are associated with substantially greater morbidity and mortality than either condition alone.^1,2^ Among patients with HF, T2D is common and independently increases the risk of cardiovascular death, HF hospitalization, and all-cause mortality.^1,3^ This risk is particularly relevant among patients with heart failure and reduced ejection fraction (HFrEF), in whom diabetes is associated with worse long-term survival despite advances in guideline-directed medical therapy (GDMT).^4^

Glucagon-like peptide-1 receptor agonists (GLP-1 RAs) have demonstrated cardiovascular benefit in large randomized trials, including reductions in major adverse cardiovascular events (MACE) and mortality.^5,6^ Their role in patients with established HFrEF; however, remains uncertain. Earlier randomized trials of liraglutide in HFrEF, including FIGHT and LIVE, failed to demonstrate clinical benefit and raised concerns regarding HF-related outcomes.^7,8^ More recent data have been more encouraging. A pre-specified analysis of the SELECT trial,^9^ which investigated the effect of semaglutide therapy on cardiovascular outcomes in patients with obesity, demonstrated cardiovascular benefit with semaglutide among patients with prevalent HF, including those with investigator-defined HFrEF, while a contemporary meta-analysis suggested lower cardiovascular mortality but no reduction in HF hospitalization among patients with HFrEF treated with GLP-1 RAs.^10,11^

GLP-1 RAs are not currently part of the foundational pharmacologic regimen for HFrEF, and their role as HF-directed therapy remains undefined.^12^ Given the conflicting evidence and limited representation of patients with established HFrEF in prior randomized trials, additional data are needed to clarify their potential role in this population. We therefore evaluated the association between GLP-1 RA use and all-cause mortality, cardiovascular events, and

HF-related outcomes among patients with T2D and HFrEF using a large propensity score-matched cohort from the TriNetX Research Network.

## METHODS

### Study Setting and Design

The current study accessed the TriNetX platform to conduct a retrospective cohort design. TriNetX’s Research Network collects longitudinal de-identified patient data from the electronic health records of 118 medical centers worldwide. Data are pooled from various healthcare encounters spanning hospital admissions, emergency department (ED) visits, outpatient clinic appointments, and telehealth clinics. Aggregated data include patient demographics (age, sex, race, ethnicity, etc.), diagnoses (defined by the International Statistical Classification of Diseases and Related Health Problems [ICD] − 10 codes), procedures (using Current Procedural Terminology [CPT] codes), laboratory results, medications prescribed, and clinical outcomes.

### Study Population

The current study collected data on adult patients, aged 18-years or older, with a diagnosis in T2D (ICD-10: E11) and a first diagnosis in HFrEF (ICD-10: I50.2 or I50.4) that occurred during the study period from May 13, 2022 (the date of US food and drug administration [FDA] approval of tirzepatide for the treatment of T2D) to July 6, 2026. The study population was divided into two study groups according to whether or not the patient had a first prescription of a GLP1-RA during the study period. The study defined GLP1-RAs as any prescription for a GLP-1 agonist (Anatomical Therapeutic Chemical [ATC] code: A10BJ) or tirzepatide. The time of study enrollment or index event was defined as the date of first GLP1-RA prescription for the GLP-1 group and the time of HFrEF diagnosis for the control group. Other exclusion criteria included the presence of medical contraindications to GLP-1 RA as detailed in the drug packaging; namely, medullary thyroid cancer (ICD-10: C73) and multiple endocrine neoplasia type 2 (ICD-10: E31.22, E31.23)

### Outcomes

Two primary outcomes were analyzed. The first was the incidence of all-cause death within one-year of follow-up. The second primary outcome was a composite of acute myocardial infarction (ICD-10 I21.9), stroke (ICD-10 I60.0─I60.3), and acute heart failure (ICD-10 I50.21, I50.23, I50.31, I50.33, I50.41, I50.43, I50.811, I50.813), defined as the first occurrence of any component of the composite outcome within one-year of follow-up. Secondary outcomes included each separate component of the primary composite outcome, and any hospital encounter (ED visit or inpatient hospitalization).

### Data collection and definitions

The current study conducted on-to-one propensity score matching of clinically relevant covariates to adjust for possible confounding factors. Covariates included in propensity score matching were sociodemographic factors (age at index, sex, race, ethnicity, socioeconomic status), comorbidities (obesity, hypertension, dyslipidemia, ischemic heart disease, atrial fibrillation or flutter, cerebrovascular disease, sleep apnea, chronic kidney disease, and chronic obstructive pulmonary disease), behavioral disorders (tobacco smoking, alcohol use disorder), body mass index, systolic blood pressure, medications (insulin, metformin, sodium-glucose cotransporter-2 [SGLT2] inhibitors, beta-blockers, renin-angiotensin-aldosterone system inhibitors, mineralocorticoid receptor antagonists, lipid-lowering medications, and antithrombotic agents), and laboratory values (low-density lipoprotein cholesterol, glycated hemoglobin [HbA1C], estimated glomerular filtration rate, and urine microalbumin-to-creatinine ratio). All covariates were assessed beginning one year prior to the index event. Covariate definitions are provided in **Table S1** of the Supplementary Materials.

### Ethics approval and informed consent

All data used in the current study were collected from the TriNetX platform, which contains de-identified patient data only per the de-identification standard defined in Section §164.514(a) of the HIPAA Privacy Rule. Thus, the current study was exempt from institutional review board (IRB) approval and informed patient consent. The study was performed in line with the principles of the Declaration of Helsinki. All data analyses are secondary analyses of existing data. The study does not involve intervention or interaction with human subjects.

### Statistical analysis

Sociodemographic and clinical characteristics were compared between the two study groups at baseline. Categorical variables were reported as numbers and percentages and continuous variables as medians and interquartile ranges (IQR). Propensity sore matching was performed by fitting logistic regression models, that were adjusted for all covariates, using greedy nearest neighbor matching with calipers of width equal to 0.1 SD of the logit of the propensity score. To assess covariate balance between the two study groups after matching, standardized mean differences (SMD) were calculated, with SMD<0.1 representing negligible covariate difference. The cumulative incidence of an outcome was calculated by dividing the number of patients in a group that had an occurrence of the outcome within the follow-up period by the number of patients in the group at the start of follow-up. Absolute risk differences (ARD) (the difference between the cumulative incidence of an outcome in the GLP-1 group and the cumulative incidence of that outcome in the control group) were calculated along with respective 95% confidence intervals (CI). Kaplan Meier curves were constructed with intergroup differences assessed using log-rank tests. Cox proportional hazards analysis was performed to calculate hazard ratios (HR) with 95% CIs.

Differences in treatment effects were estimated by performing subgroup analyses by age (65-years or older vs. Younger than 65 years), sex (male vs. Female), obesity, hypertension, CKD, sleep apnea, ischemic heart disease, cerebrovascular disease, and concomitant use of any guideline-directed medical therapy (GDMT) agent, including SGLT2 inhibitors, beta-blockers, RAAS inhibitors, or mineralocorticoid antagonists. Additional propensity score matched cohorts were built for each subgroup analysis. The difference in treatment effects between the patients who received GLP1-RA therapy and controls on the primary outcomes in each subgroup was analyzed, and the HR was calculated with the corresponding 95% CIs.

All analyses were conducted using the TriNetX platform (TriNetX LLC, Cambridge, MA, USA). A two-sided p value < 0.05 was considered statistically significant.

## RESULTS

### Study population

During the study period (May 13, 2022 to July 16, 2026), 532,581 patients had diagnoses in T2D and HFrEF in the TriNetX platform. Of these, 491,314 met the inclusion and exclusion criteria and 40,245 patients were prescribed a GLP1-RA. After propensity score matching, two cohorts with 38,879 patients in each were formed (**Fig 1**).

**Fig 1.**
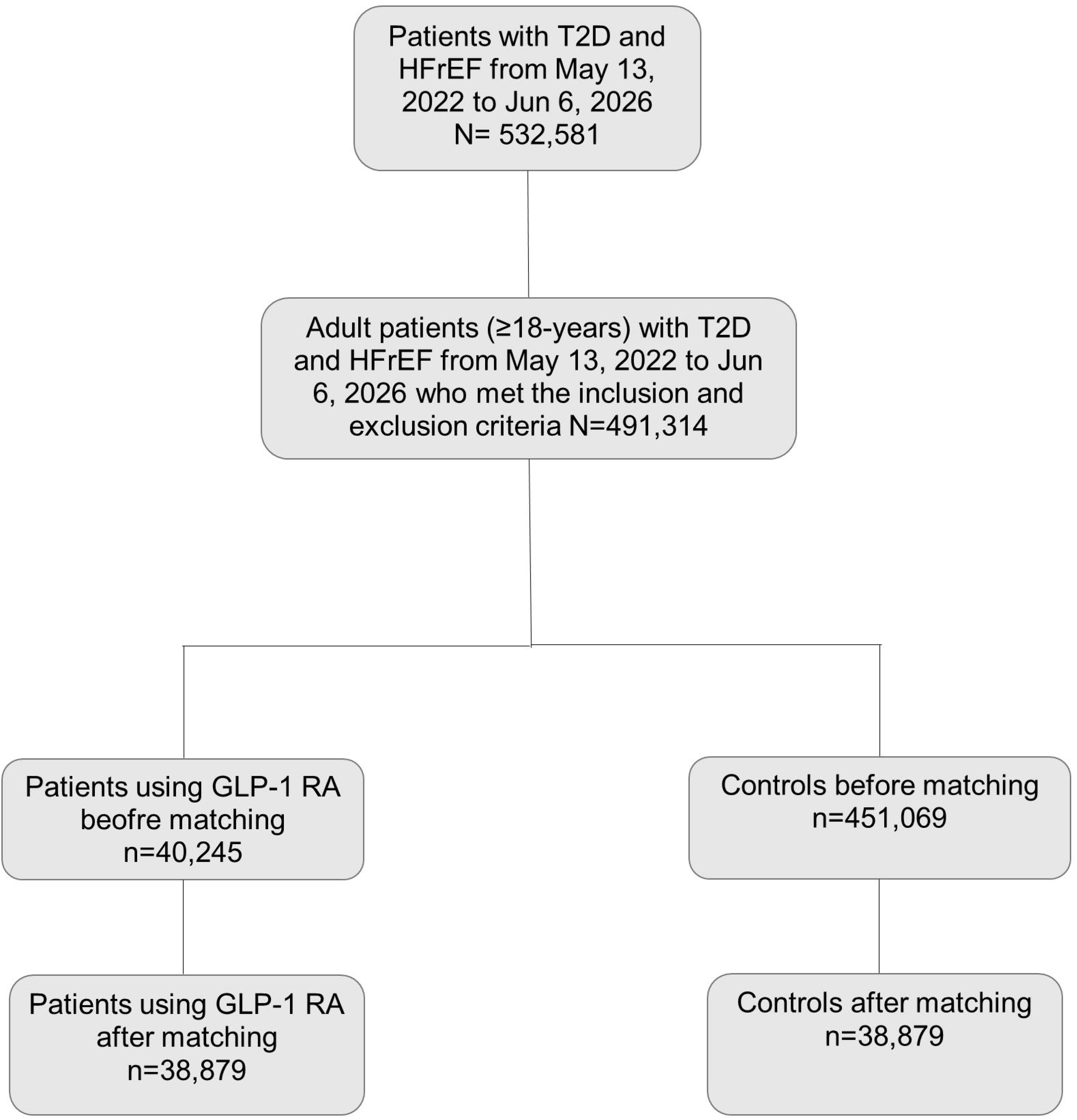
Study population flow diagram.

### Baseline clinical characteristics

The baseline sociodemographic and clinical characteristics of the two study groups before and after propensity score matching are reported in **Table 1**.

**Table 1.**
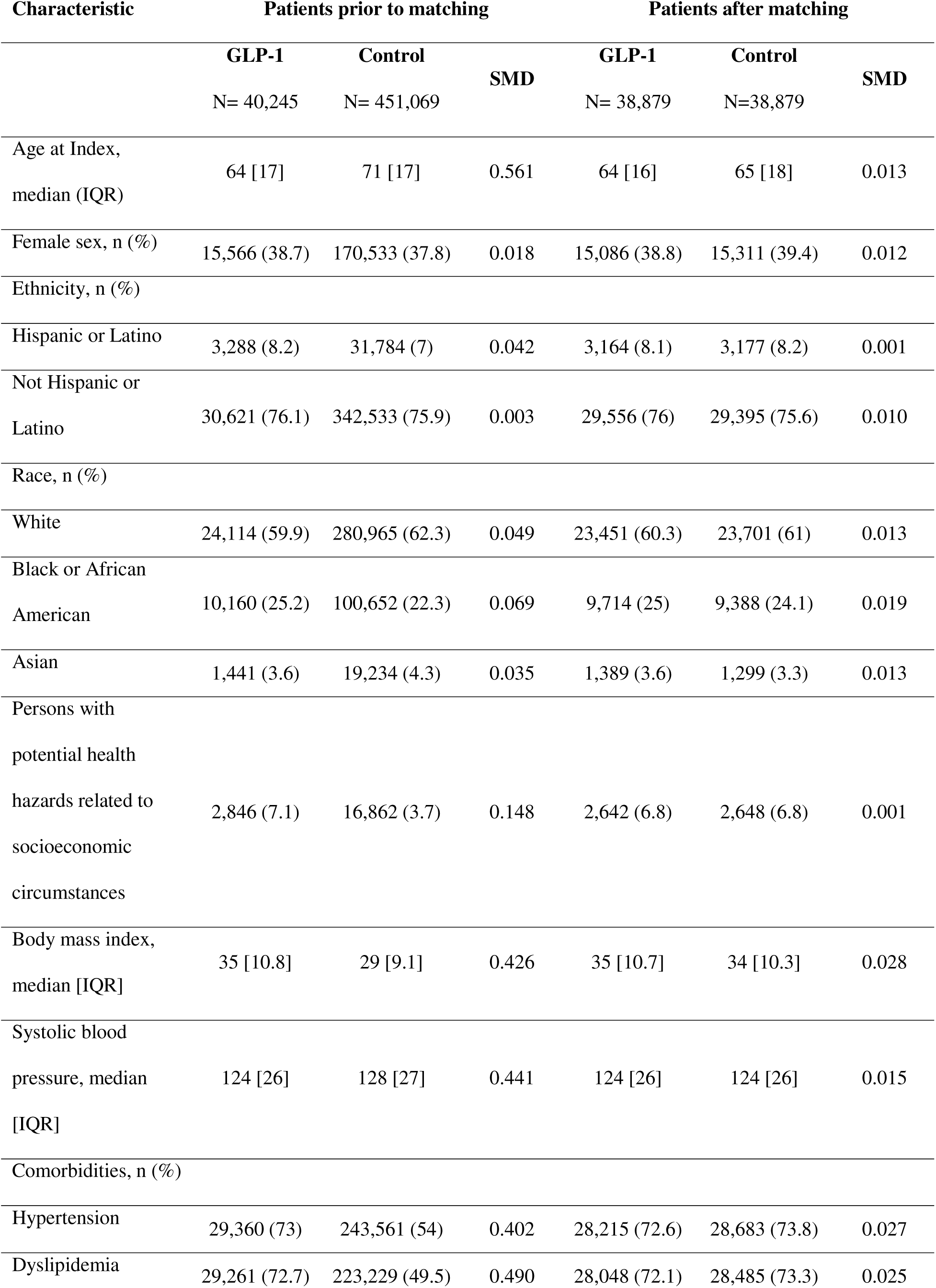
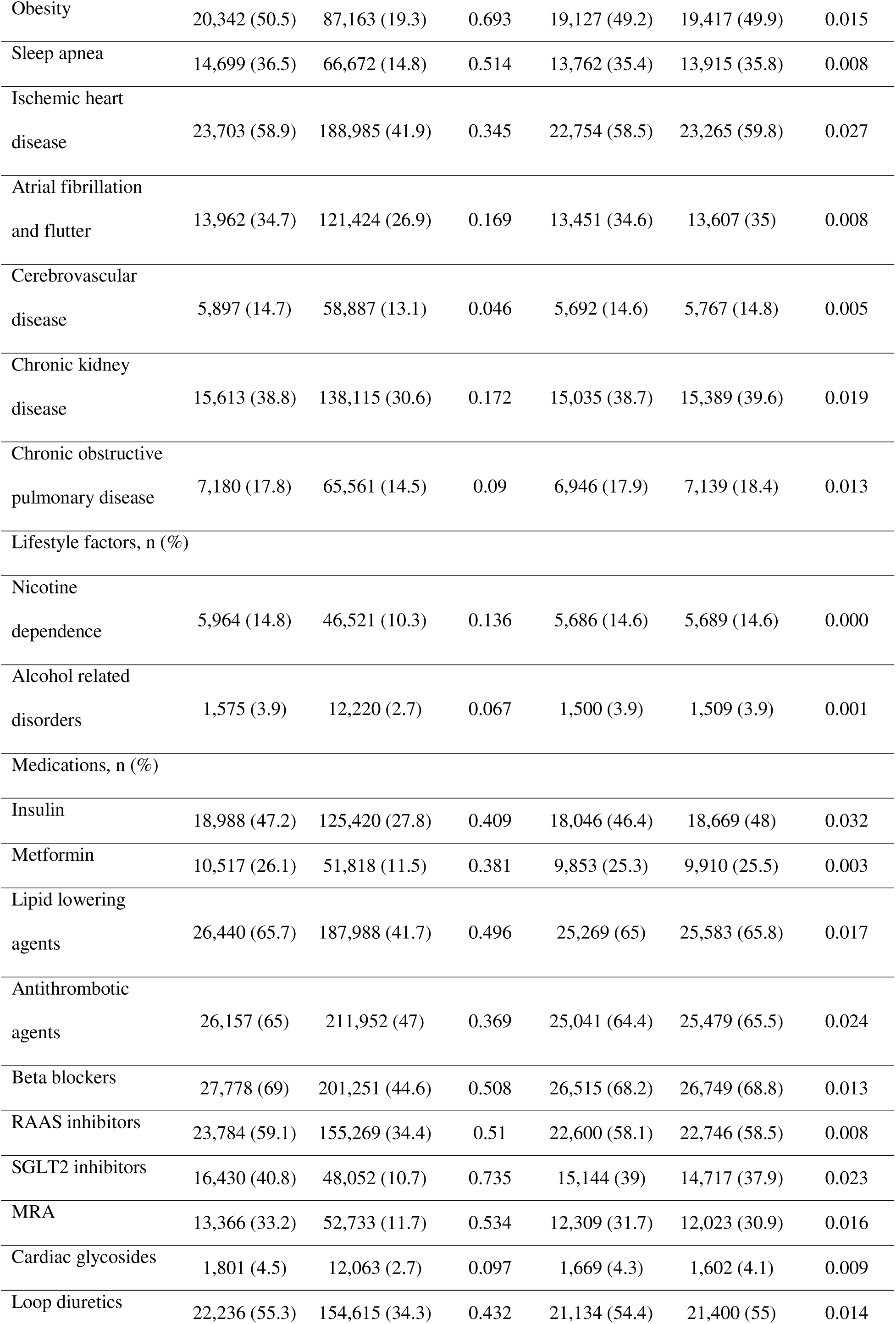
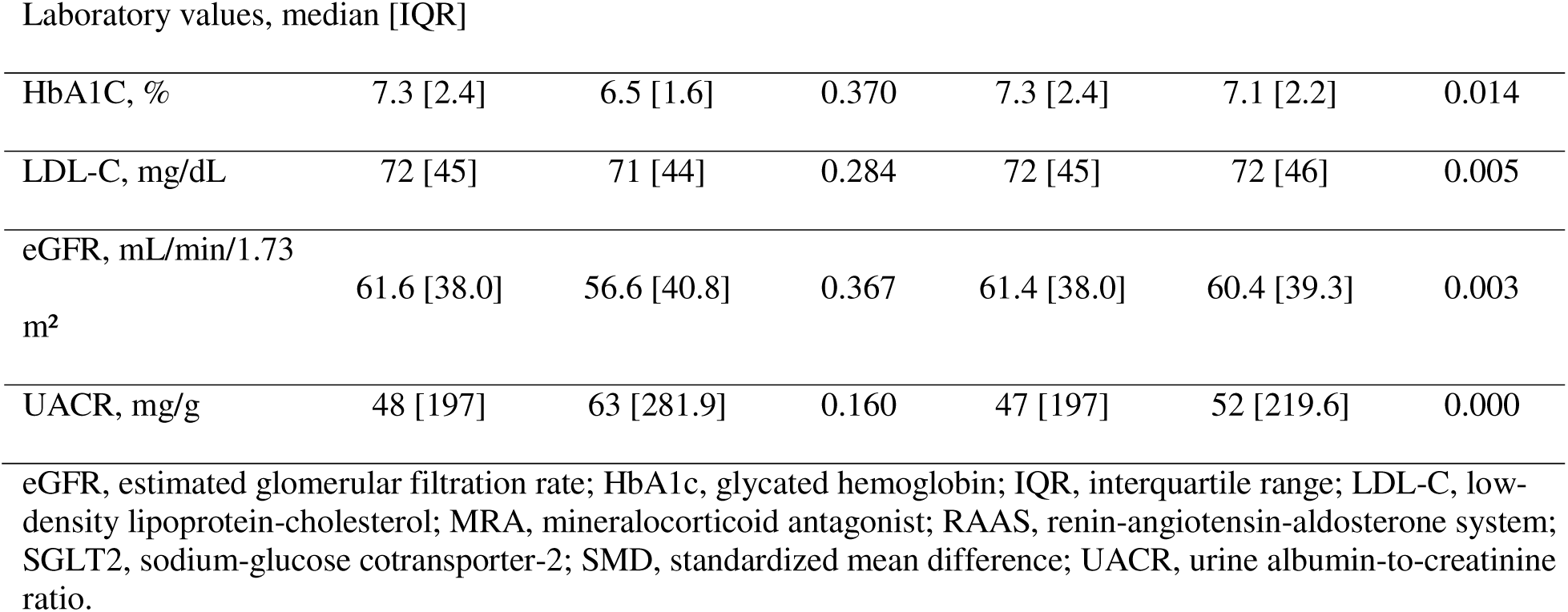
Baseline characteristics of patients with type 2 diabetes and heart failure with reduced ejection fraction stratified by use of GLP-1 receptor agonists.

| Characteristic | Patients prior to matching |  |  | Patients after matching |  |  |
| --- | --- | --- | --- | --- | --- | --- |
|  | GLP-1<br>N= 40,245 | Control<br>N= 451,069 | SMD | GLP-1<br>N= 38,879 | Control<br>N=38,879 | SMD |
| Age at Index,<br>median (IQR) | 64 [17] | 71 [17] | 0.561 | 64 [16] | 65 [18] | 0.013 |
| Female sex, n (%) | 15,566 (38.7) | 170,533 (37.8) | 0.018 | 15,086 (38.8) | 15,311 (39.4) | 0.012 |
| Ethnicity, n (%) |  |  |  |  |  |  |
| Hispanic or Latino | 3,288 (8.2) | 31,784 (7) | 0.042 | 3,164 (8.1) | 3,177 (8.2) | 0.001 |
| Not Hispanic or<br>Latino | 30,621 (76.1) | 342,533 (75.9) | 0.003 | 29,556 (76) | 29,395 (75.6) | 0.010 |
| Race, n (%) |  |  |  |  |  |  |
| White | 24,114 (59.9) | 280,965 (62.3) | 0.049 | 23,451 (60.3) | 23,701 (61) | 0.013 |
| Black or African<br>American | 10,160 (25.2) | 100,652 (22.3) | 0.069 | 9,714 (25) | 9,388 (24.1) | 0.019 |
| Asian | 1,441 (3.6) | 19,234 (4.3) | 0.035 | 1,389 (3.6) | 1,299 (3.3) | 0.013 |
| Persons with<br>potential health<br>hazards related to<br>socioeconomic<br>circumstances | 2,846 (7.1) | 16,862 (3.7) | 0.148 | 2,642 (6.8) | 2,648 (6.8) | 0.001 |
| Body mass index,<br>median [IQR] | 35 [10.8] | 29 [9.1] | 0.426 | 35 [10.7] | 34 [10.3] | 0.028 |
| Systolic blood<br>pressure, median<br>[IQR] | 124 [26] | 128 [27] | 0.441 | 124 [26] | 124 [26] | 0.015 |
| Comorbidities, n (%) |  |  |  |  |  |  |
| Hypertension | 29,360 (73) | 243,561 (54) | 0.402 | 28,215 (72.6) | 28,683 (73.8) | 0.027 |
| Dyslipidemia | 29,261 (72.7) | 223,229 (49.5) | 0.490 | 28,048 (72.1) | 28,485 (73.3) | 0.025 |
| Obesity | 20,342 (50.5) | 87,163 (19.3) | 0.693 | 19,127 (49.2) | 19,417 (49.9) | 0.015 |
| Sleep apnea | 14,699 (36.5) | 66,672 (14.8) | 0.514 | 13,762 (35.4) | 13,915 (35.8) | 0.008 |
| Ischemic heart disease | 23,703 (58.9) | 188,985 (41.9) | 0.345 | 22,754 (58.5) | 23,265 (59.8) | 0.027 |
| Atrial fibrillation and flutter | 13,962 (34.7) | 121,424 (26.9) | 0.169 | 13,451 (34.6) | 13,607 (35) | 0.008 |
| Cerebrovascular disease | 5,897 (14.7) | 58,887 (13.1) | 0.046 | 5,692 (14.6) | 5,767 (14.8) | 0.005 |
| Chronic kidney disease | 15,613 (38.8) | 138,115 (30.6) | 0.172 | 15,035 (38.7) | 15,389 (39.6) | 0.019 |
| Chronic obstructive pulmonary disease | 7,180 (17.8) | 65,561 (14.5) | 0.09 | 6,946 (17.9) | 7,139 (18.4) | 0.013 |
| Lifestyle factors, n (%) |  |  |  |  |  |  |
| Nicotine dependence | 5,964 (14.8) | 46,521 (10.3) | 0.136 | 5,686 (14.6) | 5,689 (14.6) | 0.000 |
| Alcohol related disorders | 1,575 (3.9) | 12,220 (2.7) | 0.067 | 1,500 (3.9) | 1,509 (3.9) | 0.001 |
| Medications, n (%) |  |  |  |  |  |  |
| Insulin | 18,988 (47.2) | 125,420 (27.8) | 0.409 | 18,046 (46.4) | 18,669 (48) | 0.032 |
| Metformin | 10,517 (26.1) | 51,818 (11.5) | 0.381 | 9,853 (25.3) | 9,910 (25.5) | 0.003 |
| Lipid lowering agents | 26,440 (65.7) | 187,988 (41.7) | 0.496 | 25,269 (65) | 25,583 (65.8) | 0.017 |
| Antithrombotic agents | 26,157 (65) | 211,952 (47) | 0.369 | 25,041 (64.4) | 25,479 (65.5) | 0.024 |
| Beta blockers | 27,778 (69) | 201,251 (44.6) | 0.508 | 26,515 (68.2) | 26,749 (68.8) | 0.013 |
| RAAS inhibitors | 23,784 (59.1) | 155,269 (34.4) | 0.51 | 22,600 (58.1) | 22,746 (58.5) | 0.008 |
| SGLT2 inhibitors | 16,430 (40.8) | 48,052 (10.7) | 0.735 | 15,144 (39) | 14,717 (37.9) | 0.023 |
| MRA | 13,366 (33.2) | 52,733 (11.7) | 0.534 | 12,309 (31.7) | 12,023 (30.9) | 0.016 |
| Cardiac glycosides | 1,801 (4.5) | 12,063 (2.7) | 0.097 | 1,669 (4.3) | 1,602 (4.1) | 0.009 |
| Loop diuretics | 22,236 (55.3) | 154,615 (34.3) | 0.432 | 21,134 (54.4) | 21,400 (55) | 0.014 |
| Laboratory values, median [IQR] |  |  |  |  |  |  |
| HbA1C, % | 7.3 [2.4] | 6.5 [1.6] | 0.370 | 7.3 [2.4] | 7.1 [2.2] | 0.014 |
| LDL-C, mg/dL | 72 [45] | 71 [44] | 0.284 | 72 [45] | 72 [46] | 0.005 |
| eGFR, mL/min/1.73<br>m<sup>2</sup> | 61.6 [38.0] | 56.6 [40.8] | 0.367 | 61.4 [38.0] | 60.4 [39.3] | 0.003 |
| UACR, mg/g | 48 [197] | 63 [281.9] | 0.160 | 47 [197] | 52 [219.6] | 0.000 |

eGFR, estimated glomerular filtration rate; HbA1c, glycated hemoglobin; IQR, interquartile range; LDL-C, low-density lipoprotein-cholesterol; MRA, mineralocorticoid antagonist; RAAS, renin-angiotensin-aldosterone system; SGLT2, sodium-glucose cotransporter-2; SMD, standardized mean difference; UACR, urine albumin-to-creatinine ratio.

In brief, prior to propensity score matching, the GLP-1 group were younger, had poorer socioeconomic status and greater comorbidity burden. After propensity score matching, both groups had similar characteristics with SMD <0.1 for all covariates.

### Follow-up time

The median [interquartile range] follow-up time was 365 [173] days in the GLP-1 group and 365 [119] days in the control group. The majority of patients in both groups completed one year of follow-up (**Fig 2**).

**Fig 2.**
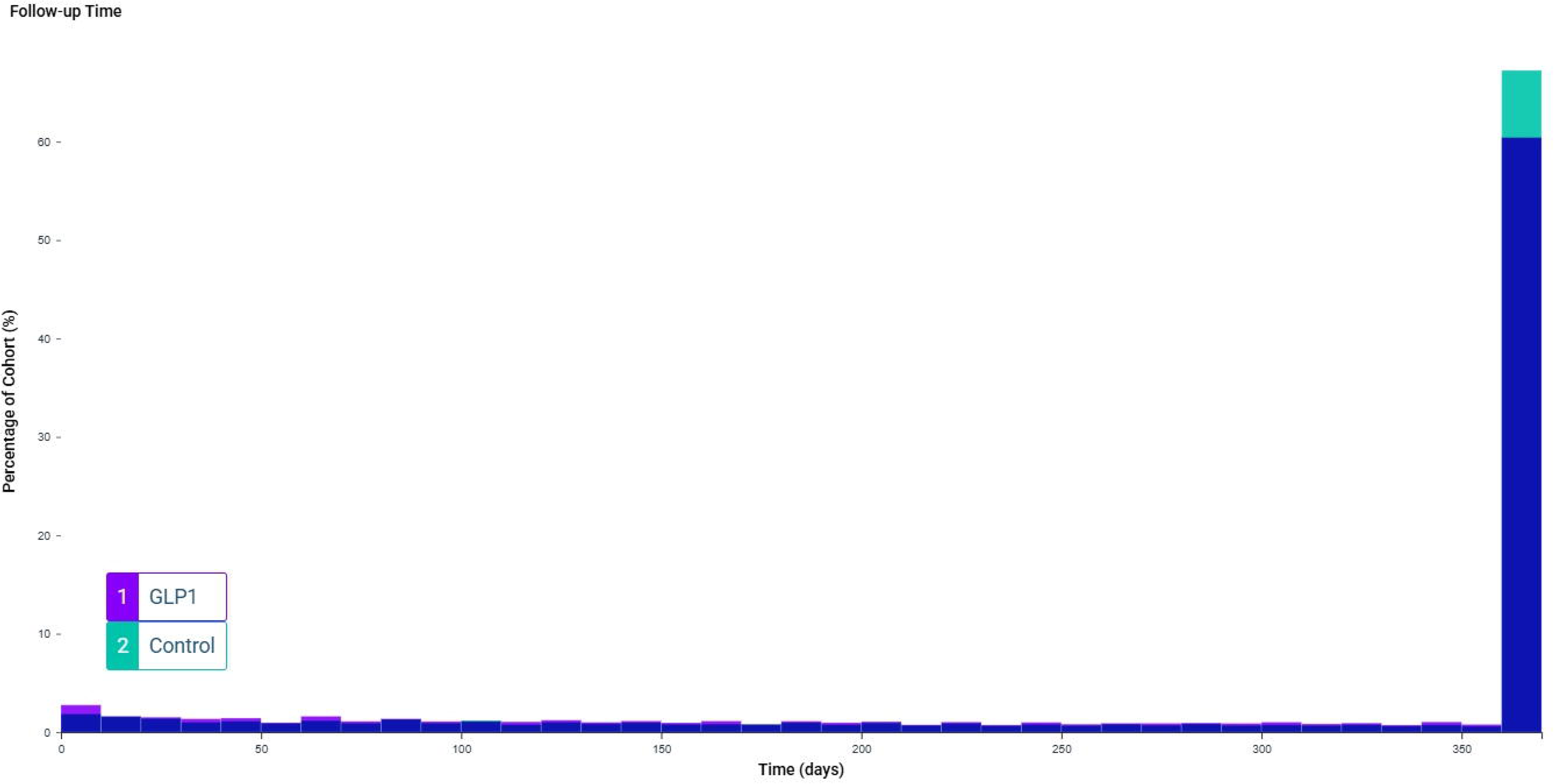
Follow-up time for patients with type 2 diabetes and heart failure with reduced ejection fraction treated with GLP-1 receptor therapy and matched controls.

### Clinical outcomes

The incidence, ARD and HR with 95% CIs for the primary and secondary outcomes are reported in **Table 2**. The GLP-1 group had lower incidences of all-cause mortality and the primary composite outcome compared to the control group (**Fig 3**). The GLP-1 group were also at lower risk of all individual components of the primary composite outcome, and any ED visit or hospitalization compared to the control group.

**Fig 3.**
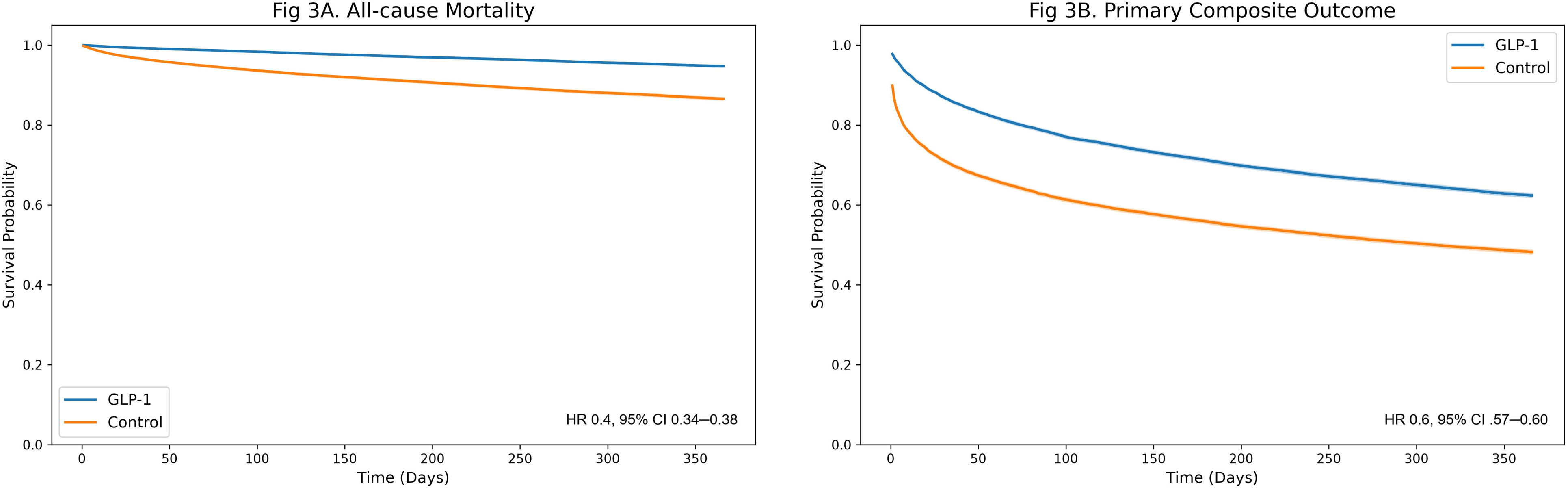
Kaplan-Meier curves comparing the incidences of all-cause mortality and a primary composite outcome in patients with type 2 diabetes and heart failure with reduced ejection fraction treated with GLP-1 receptor therapy and matched controls. The online tool, TriNetX Publication Toolkit (URL: https://trinetxpublicationtoolkit.streamlit.app/) was accessed to generate Kaplan–Meier curves.

**Table 2.** Cardiovascular outcomes in patients with heart failure and reduced ejection fraction and type 2 diabetes using GLP-1 RA.

| Outcome | No. of patients<br>with event | Cumulative<br>incidence (%) | ARD (95% CI)<br>% | HR (95% CI) |
| --- | --- | --- | --- | --- |
| **All-cause death** |  |  |  |  |
| GLP-1 RA | 1631 | 4.2 | -7.8 (-8.17 to -7.41) | 0.4 (0.34–0.38) |
| Control | 4660 | 12.0 | Ref (0) | Ref (1) |
| **Primary composite outcome** |  |  |  |  |
| GLP-1 RA | 12647 | 32.5 | -15.8 (-16.47 to -15.11) | 0.6 (0.57–0.60) |
| Control | 18785 | 48.3 | Ref (0) | Ref (1) |
| **Myocardial infarction** |  |  |  |  |
| GLP-1 RA | 4036 | 10.4 | -6.8 (-7.29 to -6.33) | 0.6 (0.57–0.62) |
| Control | 6683 | 17.2 | Ref (0) | Ref (1) |
| **Ischemic stroke** |  |  |  |  |
| GLP-1 RA | 2961 | 7.6 | -2.3 (-2.7 to -1.9) | 0.8 (0.74–0.82) |
| Control | 3856 | 9.9 | Ref (0) | Ref (1) |
| **Acute heart failure** |  |  |  |  |
| GLP-1 RA | 8894 | 22.9 | -14.4 (-14.99 to -13.72) | 0.6 (0.54–0.57) |
| Control | 14474 | 37.2 | Ref (0) | Ref (1) |
| **ED visits or hospitalizations** |  |  |  |  |
| GLP-1 RA | 16371 | 42.1 | -17.4 (-18.12 to -16.73) | 0.6 (0.57–0.60) |
| Control | 23146 | 59.5 | Ref (0) | Ref (1) |

### Subgroup analysis

GLP1-RA therapy was associated with lower incidence of all-cause mortality and the primary composite outcome compared to control across all studied sub-groups (**Fig 4**).

**Fig 4.**
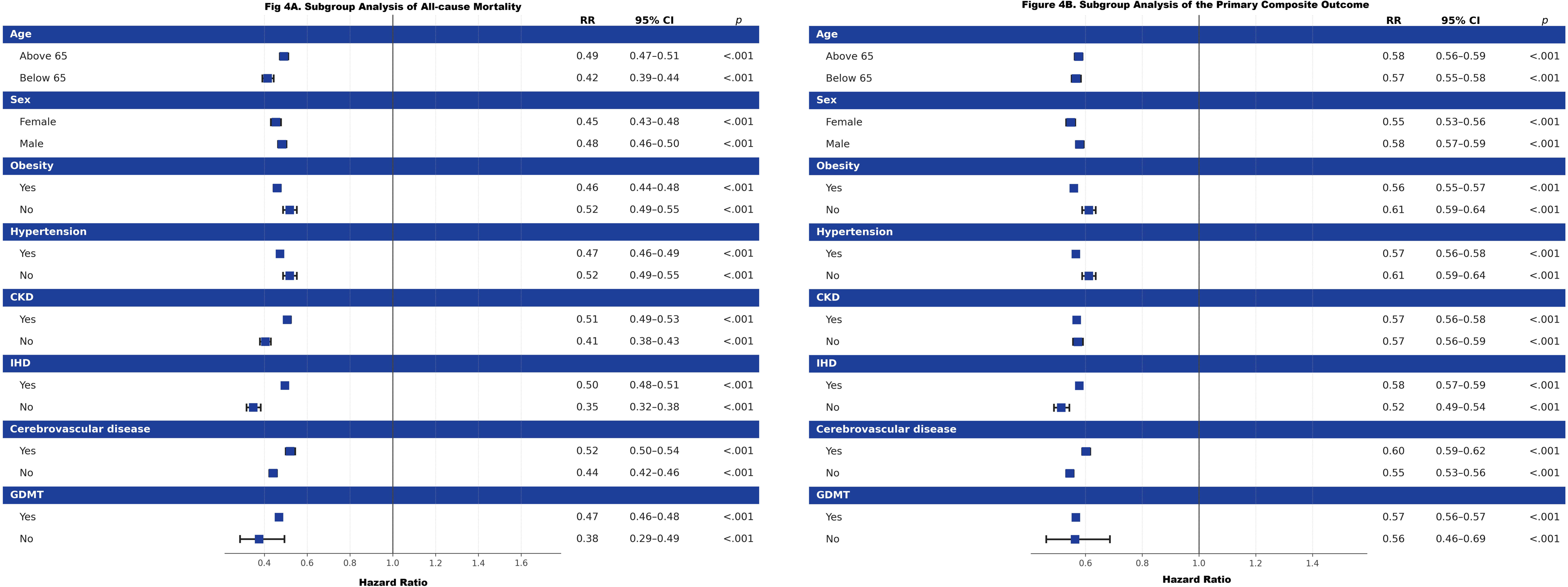
Forest plots presenting hazard ratios with 95% confidence intervals for all-cause mortality and a primary composite outcome in subgroups of patients with type 2 diabetes and heart failure with reduced ejection fraction treated with GLP-1 receptor therapy and matched controls (reference). The online tool, TriNetX Publication Toolkit (URL: https://trinetxpublicationtoolkit.streamlit.app/) was accessed to generate forest plots.

## DISCUSSION

In this large propensity score-matched cohort of patients with T2D and HFrEF, GLP-1 RA use was associated with lower rates of all-cause mortality, acute myocardial infarction, stroke, acute HF events, and the composite cardiovascular outcome compared with matched controls. Similar associations were observed across clinically relevant subgroups, including patients receiving concomitant HFrEF therapies. These findings add to a growing, but still evolving, body of evidence regarding GLP-1 RA use in patients with HFrEF.

The lower mortality observed in this study is directionally consistent with the broader cardiovascular literature on GLP-1 RAs. In a meta-analysis of 11 cardiovascular outcome trials, Kang et al. found that GLP-1 RAs reduced all-cause, cardiovascular, and noncardiovascular mortality.^13^ More recently, a meta-analysis of 21 randomized trials, including 99,599 participants, found evidence for reductions in all-cause mortality, cardiovascular mortality, and MACE with GLP-1 RA therapy.^5^ These trials, however, were not designed specifically to evaluate patients with established HFrEF. The present study extends this literature by examining a large real-world cohort specifically composed of patients with both T2D and HFrEF.

Several mechanisms could contribute to the cardiovascular associations observed in this study. GLP-1 RAs have effects extending beyond glycemic control, including reductions in body weight, blood pressure, postprandial lipemia, and systemic inflammation.^6^ Experimental and clinical data also suggest favorable effects on vascular inflammation and atherosclerotic disease progression.^14^ These pathways provide a plausible basis for reductions in atherosclerotic cardiovascular events. They do not, however, establish a direct beneficial effect on the failing myocardium, and whether GLP-1 RAs improve HFrEF-specific outcomes independently of broader cardiometabolic risk modification should be further explored in future studies.

This distinction is particularly important given the results of earlier randomized trials in HFrEF. FIGHT enrolled patients with reduced ejection fraction following a recent HF hospitalization and found no improvement in post-hospitalization clinical stability with liraglutide.^7^ Similarly, LIVE found no improvement in left ventricular systolic function and reported a greater number of serious cardiac events among patients receiving liraglutide.^8^ These studies differed considerably from the present analysis. FIGHT enrolled a particularly high-risk population following recent HF hospitalization, whereas LIVE evaluated patients with stable chronic HF. Both trials also evaluated liraglutide rather than the newer incretin-based agents that account for a growing proportion of contemporary use. Differences in patient population, disease severity, drug exposure, and study design may partly explain the differing results. At the same time, these randomized data argue against interpreting the associations observed in our study as evidence of a direct or uniform HFrEF treatment effect.

More recent evidence has been somewhat more reassuring. In a prespecified analysis of SELECT, semaglutide reduced MACE among participants with prevalent HF, with similar benefit across HF subtypes. Among participants with investigator-defined HFrEF, the HR for MACE was 0.65 (95% CI, 0.49–0.87).^10^ Important differences remain however. SELECT enrolled patients with overweight or obesity and established cardiovascular disease but without diabetes, excluded patients with New York Heart Association (NYHA) class IV HF. Its population therefore may differ from the T2D and HFrEF population examined in the present study.

The meta-analysis by Siddiqi et al. further illustrates the uncertainty surrounding HF-specific outcomes. Among patients with HFrEF, GLP-1 RA therapy was associated with lower cardiovascular mortality, but a numerically higher, nonsignificant risk of HF hospitalization.^11^ This contrasts with the lower rate of acute HF events observed in our cohort. Differences in study design, patient selection, HF severity, exposure definitions, and outcome ascertainment may account for some of this discrepancy. The discordance also underscores the need for caution in interpreting lower acute HF event rates in an observational analysis as evidence of a direct HF benefit. The findings among patients receiving concomitant GDMT are also noteworthy. In another TriNetX analysis, Kokura et al. evaluated patients with HFrEF and T2D receiving comprehensive GDMT and found lower all-cause mortality among those also receiving GLP-1 RAs, although hospitalization was not significantly reduced.^15^ In addition, a meta-analysis of randomized trials found that the cardiovascular and kidney benefits of GLP-1 RAs in T2D were consistent irrespective of baseline sodium-glucose cotransporter-2 (SGLT2) inhibitor use.^16^ Taken together, these data raise the possibility that GLP-1 RAs may provide additional cardiometabolic benefit in appropriately selected patients already receiving contemporary therapy. They do not establish GLP-1 RAs as a component of HFrEF-directed GDMT, and prospective studies are needed before such a role can be defined.

### Limitations

This study has several strengths. The large sample size allowed us to examine a clinically important population that has been incompletely represented in randomized GLP-1 RA trials and to assess the consistency of the observed associations across multiple clinically relevant subgroups. Propensity score matching incorporated a broad range of demographic characteristics, comorbidities, medications, vital signs, and laboratory measurements, with standardized mean differences <0.1 after matching for all included covariates.

Several limitations should be considered when interpreting these findings. The retrospective observational design precludes causal inference. Propensity score matching can reduce imbalance in measured characteristics but cannot account for unmeasured or incompletely measured confounders. Patients prescribed GLP-1 RAs may also differ systematically from untreated patients in healthcare engagement, access to care, weight-management efforts, and other factors that influence outcomes.

The use of electronic health record data introduces additional potential for misclassification and incomplete outcome capture. Diagnoses and outcomes depend on coding within participating healthcare organizations, and medication prescriptions do not establish that a medication was dispensed or taken. Deaths occurring outside participating healthcare organizations may also be incompletely captured in electronic health record data, potentially resulting in misclassification of vital status.^17^ The study is also open to biases inherent to observational studies.

## CONCLUSION

GLP-1 RA use was associated with lower all-cause mortality and fewer cardiovascular and acute HF events among patients with T2D and HFrEF in this large propensity score-matched real-world cohort. Randomized trials with rigorous HF phenotyping are needed to determine whether GLP-1 RAs directly improve clinical outcomes in patients with established HFrEF and to define their role alongside contemporary GDMT.

## Data Availability

All data produced are available online at (https://live.trinetx.com/).

## Statements and Declarations

### Ethics approval

This study was performed in line with the principles of the Declaration of Helsinki. All data used were collected from the TriNetX platform, which contains de-identified patient data only. Thus, an institutional review board (IRB) approval and patient consent were not required.

### Consent to participate declaration

The current study is exempt from informed consent. All data analyses are secondary analyses of existing data. The study does not involve intervention or interaction with human subjects. All included data are de-identified per the de-identification standard defined in Section §164.514(a) of the HIPAA Privacy Rule.

### Funding declaration

This research did not receive any specific grant from funding agencies in the public, commercial, or not-for-profit sectors.

Declaration of generative AI and AI-assisted technologies in the manuscript preparation process.

During the preparation of this work the authors used the online tool, TriNetX Publication Toolkit (URL: https://trinetxpublicationtoolkit.streamlit.app/) in order to generate Kaplan–Meier curves and forest plots. After using this tool/service, the authors reviewed and edited the content as needed and takes full responsibility for the content of the published article.

## References

1. Dunlay SM, Givertz MM, Aguilar D, et al. Type 2 Diabetes Mellitus and Heart Failure: A Scientific Statement From the American Heart Association and the Heart Failure Society of America: This statement does not represent an update of the 2017 ACC/AHA/HFSA heart failure guideline update. Circulation. 2019;140(7):e294–e324. doi:10.1161/CIR.0000000000000691

2. Seferović PM, Petrie MC, Filippatos GS, et al. Type 2 diabetes mellitus and heart failure: a position statement from the Heart Failure Association of the European Society of Cardiology. Eur J Heart Fail. 2018;20(5):853–872. doi:10.1002/ejhf.1170

3. Dauriz M, Mantovani A, Bonapace S, et al. Prognostic Impact of Diabetes on Long-term Survival Outcomes in Patients With Heart Failure: A Meta-analysis. Diabetes Care. 2017;40(11):1597–1605. doi:10.2337/dc17-0697

4. Fudim M, Devaraj S, Chukwurah M, et al. Prognosis for patients with heart failure and reduced ejection fraction with and without diabetes: A 7 year nationwide veteran administration analysis. Int J Cardiol. 2022;346:30–34. doi:10.1016/j.ijcard.2021.11.032

5. Galli M, Benenati S, Laudani C, et al. Cardiovascular Effects and Tolerability of GLP-1 Receptor Agonists: A Systematic Review and Meta-Analysis of 99,599 Patients. J Am Coll Cardiol. 2025;86(20):1805–1819. doi:10.1016/j.jacc.2025.08.027

6. Ussher JR, Drucker DJ. Glucagon-like peptide 1 receptor agonists: cardiovascular benefits and mechanisms of action. Nat Rev Cardiol. 2023;20(7):463–474. doi:10.1038/s41569-023-00849-3

7. Margulies KB, Hernandez AF, Redfield MM, et al. Effects of Liraglutide on Clinical Stability Among Patients With Advanced Heart Failure and Reduced Ejection Fraction: A Randomized Clinical Trial. JAMA. 2016;316(5):500–508. doi:10.1001/jama.2016.10260

8. Jorsal A, Kistorp C, Holmager P, et al. Effect of liraglutide, a glucagon-like peptide-1 analogue, on left ventricular function in stable chronic heart failure patients with and without diabetes (LIVE)-a multicentre, double-blind, randomised, placebo-controlled trial. Eur J Heart Fail. 2017;19(1):69–77. doi:10.1002/ejhf.657

9. Lincoff AM, Brown-Frandsen K, Colhoun HM, et al. Semaglutide and Cardiovascular Outcomes in Obesity without Diabetes. N Engl J Med. 2023;389(24):2221–2232. doi:10.1056/NEJMoa2307563

10. Deanfield J, Verma S, Scirica BM, et al. Semaglutide and cardiovascular outcomes in patients with obesity and prevalent heart failure: a prespecified analysis of the SELECT trial. *Lancet (London*, England). 2024;404(10454):773–786. doi:10.1016/S0140-6736(24)01498-3

11. Siddiqi TJ, Khan MS, Waqas SA, et al. Effect of glucagon-like peptide-1 receptor agonists on heart failure outcomes and cardiovascular death across varying cardiovascular-kidney-metabolic comorbidity. Eur J Heart Fail. 2025;27(12):2844–2854. doi:10.1002/ejhf.70048

12. Heidenreich PA, Bozkurt B, Aguilar D, et al. 2022 AHA/ACC/HFSA Guideline for the Management of Heart Failure: A Report of the American College of Cardiology/American Heart Association Joint Committee on Clinical Practice Guidelines. Circulation. 2022;145(18):e895-e1032. doi:10.1161/CIR.0000000000001063

13. Kang YM, Bohula EA, Lincoff AM, et al. Glucagon-like peptide-1 receptor agonists lower risk of cardiovascular and non-cardiovascular mortality: A meta-analysis of eleven cardiovascular outcome trials. Diabetes Obes Metab. 2025;27(7):4017–4021. doi:10.1111/dom.16424

14. Kassab MB, Khraishah H, Thrapp A, et al. Glucagon-like peptide-1 receptor agonists reduce experimental atherosclerosis progression, inflammatory biomarkers and cardiovascular events, irrespective of hyperglycaemia and obesity. Eur Heart J. 2026;47(28):3800–3817. doi:10.1093/eurheartj/ehag060

15. Kokura Y, Kishimori T, Kato T, et al. Impact of glucagon-like peptide-1 receptor agonists with guideline-directed medical therapy in patients with heart failure with reduced ejection fraction and type 2 diabetes. Diabetes Res Clin Pract. 2026;238:113404. 10.1016/j.diabres.2026.113404

16. Neuen BL, Fletcher RA, Heath L, et al. Cardiovascular, Kidney, and Safety Outcomes With GLP-1 Receptor Agonists Alone and in Combination With SGLT2 Inhibitors in Type 2 Diabetes: A Systematic Review and Meta-Analysis. Circulation. 2024;150(22):1781–1790. doi:10.1161/CIRCULATIONAHA.124.071689

17. Olaker VR, Fry S, Terebuh P, et al. With big data comes big responsibility: Strategies for utilizing aggregated, standardized, de-identified electronic health record data for research. Clin Transl Sci. 2025;18(1):e70093. doi:10.1111/cts.70093

