## Supplementary Materials for "Cardiovascular Outcomes in Patients with Type 2 Diabetes and Heart Failure with Reduced Ejection using Glucagon-like Peptide-1 Receptor Agonists"

Affiliations:
^1^Department of Internal Medicine, Georgetown University- MedStar Washington Hospital Center, 110 Irving St NW, Washington, DC 20010, USA.

^2^Department of Internal Medicine, MedStar Georgetown University Hospital, 3800 Reservoir Rd NW, Washington, DC 20007, USA.
^3^Department of Internal Medicine, USF Health Morsani College of Medicine, 13220 USF Laurel Dr., Tampa, FL 33612, USA.

**Table S1**. Covariate definitions.

| **Covariate** | **Definition (ICD-10 Code)** |
| --- | --- |
| Socioeconomic status | Z55-Z65 |
| Overweight or obesity | E66 |
| hypertension | I10 |
| dyslipidemia | E78 |
| ischemic heart disease | I20-I25 |
| atrial fibrillation or flutter | I48 |
| cerebrovascular disease | I60-I69 |
| sleep apnea | G47.3 |
| Chronic kidney disease | N18 |
| chronic obstructive pulmonary disease | J44 |
| tobacco smoking | F17 |
| alcohol use disorder | F10 |
| Insulin | ATC- A10A |
| Metformin | RXCUI- 6809 |
| SGLT2 inhibitors | ATC- A10BK |
| Lipid-lowering agents | ATC- C10 |
| Beta-blockers | ATC- C07 |
| RAAS inhibitors | ATC- C09 |
| Mineralocorticoid antagonists | ATC- C03DA |
| Antithrombotics | ATC- B01 |
| Cardiac glycosides | ATC- C01A |
| Loop diuretics | ATC- C03CA |
